# Preliminary Estimates of Years of Life Lost (YLL) Due to COVID-19 in India

**DOI:** 10.1101/2020.10.24.20218693

**Authors:** Abhaya Indrayan, Aman Mishra, Mona Pathak

## Abstract

**Objectives:** More than 7 million cases of COVID have been detected in India by the middle of October 2020 and more than 100 thousand deaths have occurred. In this communication, we present an estimate of the years of life lost (YLL) due to COVID-19 so far and the projection for the full year so that the health damage by this new disease can be compared with some other ailments.

**Design:** Records based study

**Method:** The YLL by one premature death is the expectation of life at that age. To calculate YLL, the age-wise distribution of COVID cases and deaths was obtained from the official sources of the government of India. Similar calculations were done for the general population from all causes for comparison.

**Results:** A total of more than 2 million years of life have already lost due to COVID-19 and the pattern indicates that we may end up with nearly 4 million YLL due to this disease in India. This is nearly 20 years lost per COVID death, 303 years lost per 1000 cases of COVID, and about 3 years lost per 1000 population in a full year. The age-group 50–59 years has been particularly affected. Other important findings are summarized as key messages.

**Conclusion:** The years of life lost so far and anticipated in full-year are enormous but may still be lower compared with some other causes such as road injuries.

## Introduction

COVID-19 has caused havoc around the world and India seems to be going to head the table as a country with the highest number of cases. We are already second highest in the middle of October 2020 but the cases are rising faster than any other country. More than 7.2 million cases have been reported and more than 110 thousand deaths are attributed to this disease during the past 7 months when it started reaching epidemic proportion. The mortality rate was 1.52% in the middle of October 2020. But the years of life lost (YLL) due to COVID in India is not yet known.

The YLL for a person is how long the person would have lived had not died prematurely. This metric is crucial to understand the full impact of a disease on the longevity of the people. YLL calculation at macro level accounts not just how much life is cut short for the persons who succumbed but also the number of deaths at different ages.

This paper presents the estimates of the YLL per COVID death, per 1000 COVID cases, and per thousand population. Because of limitation of the availability of age wise data on cases and deaths, and because the epidemic is still going on, the estimates are preliminary, although they may be enough to provide a lead to future efforts in this direction.

## Material & Methods

Although the country wide information on age-wise incidence of COVID and deaths is meagre in India, some information has recently come in the public domain. The distribution of COVID cases in 10-year age intervals was extracted from the website of National Centre for Disease Control (NCDC) [1] of the government of India. The latest information as of October 17, 2020, was available for 2,878,619 cases and this sample is large enough to be a fair representation of age distribution of all 7.4 million cases occurred till October 17, 2020, as the epidemic has stabilized by that time for age-wise incidence. This is the cut-off date for our data. The age distribution of COVID deaths was obtained from a newspaper dated September 2, 2020 [2] who also reported NCDC as the source. This was available for 56,288 deaths till that date, which also seems a large enough number to provide the age pattern of COVID deaths in India. A subsequent article in The Wire on October 14, 2020, quoting the government sources, also gave nearly the same age distribution of more than 110 thousand deaths of COVID by that time [3].

The calculation of YLL is based on the expectation of life in India at the mid-point of the age-groups at death. For example, if a person of age-group 50–59 years dies, the expectation of life at age 55 years is 21.8 years, and this is lost due to this death. This is the YLL due to premature death at that age. The expectation of life at different ages was taken from the Abridged Life Tables of the Sample Registration System for the years 2014-18 [4]. This is the latest available (2020 publication) and may not have changed much since then. Thus, our calculations for YLL are based on the latest available Indian life expectancy and not based on any fixed life expectancy or the highest life expectancy used by international agencies [5]. For this reason, our estimates are not internationally comparable but are rooted to the Indian realities and the current situation. No disaggregation was done by gender as the breakup of COVID cases and deaths by gender (in different age-groups) was not available.

The calculation of YLL per 1000 population was done as follows:

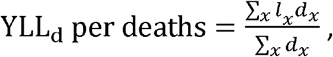

where, *l*_*x*_ = expectation of life at age *x*, and

*d*_*x*_ = number of deaths by COVID at age *x*.

If the mortality rate in COVID cases is M%, the YLL per thousand cases will be:

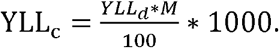

If the incidence of the disease is I per 1000 population, the YLL will be:

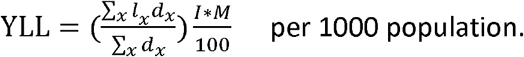

All calculations were done for the mid-point of each 10-year interval. No age weighing or discounting was used as per the latest methodology [5].

A similar exercise was also done for the general population of India using the age distribution of population and deaths reported by the latest (2018) report of the Sample Registration System [6].

Since very few COVID deaths occurred before the age of 20 years, whereas these are far more in the general population due to high infant and child mortality in India, we separately report YLL for the age 20 years and above.

## Results

Basic data on the age distribution of cases and deaths by COVID and the general population (Table I) show that only about 12% COVID cases in India had age less than 20 years whereas the share of this age-group was nearly 36% in the general population. Thus, the relative incidence of COVID in India is just about one-third in this age-group. Compare this with nearly 12% COVID cases of age 60 years and above vs. 8% population in this age-group. Thus, the age-group 60 years and above was disproportionately more affected. But the incidence of COVID has been particularly disproportionately higher in the age-group 50-59 years with a share of 14.5% of COVID cases compared to only 7.7% share of this age-group in the population. Thus this age-group was twice as commonly affected. The share (0.9%) of age-group 80 years and above in COVID cases was nearly the same as in the general population (0.8%).

**Table 1:** Age distribution of COVID cases and deaths, and the general population

| Age group (years) | COVID (% distribution) |  | General Population (% distribution) |  |
| --- | --- | --- | --- | --- |
|  | Cases | Deaths | Population | Deaths |
| 0-9 | 3.9 | 0.5 | 16.6 | 12.2 |
| 10-19 | 8.0 | 0.7 | 19.5 | 2.1 |
| 20-29 | 22.3 | 2.6 | 20.9 | 3.9 |
| 30-39 | 21.6 | 6.1 | 15.7 | 5.1 |
| 40-49 | 17.3 | 13.4 | 11.5 | 7.7 |
| 50-59 | 14.5 | 25.3 | 7.7 | 12.9 |
| 60-69 | 8.2 | 28.6 | 5.1 | 20.1 |
| 70-79 | 3.2 | 17.0 | 2.3 | 20.2 |
| >=80 | 0.9 | 5.8 | 0.8 | 15.7 |
| Total | 100 | 100 | 100 | 100 |

Less than 2% of all COVID deaths were in the age-group less than 20 years. Against this, in the total population, nearly 14% of all deaths in India were in this age-group. Thus, the disease is nearly one-seventh as fatal in this age-group as the all-cause mortality in India. The situation in the age-group 60 years and above is nearly the reverse. The share of this age-group in total COVID deaths was more than 51% whereas in the general population this was 56%. Only 5.8% of all COVID deaths were in the age-group 80 years and above while in the general population, the contribution of this age-group was 15.7%. The onslaught of mortality was particularly more in the age-group 50–59 years that accounted for 25.3% share of COVID deaths against only 12.9% share in deaths in the general population. For visual comparison of these percentages in different age groups, see Figures 1S and 2S in the Supplement.

The age wise pattern of deaths shows that the mortality rate was higher in COVID cases of age-group less than 80 years than in the general population but lower in the cases of age 80 years and above (Figure 1). This was particularly so for people of age 40–69 years where deaths in COVID cases were almost twice as common. Age-specific death rate in age-group 80+ was only 102.5 per 1000 COVID cases whereas this was 141.9 per 1000 persons in the general population. Thus, COVID cases have less death rate in this age-group.

**Figure 1.**
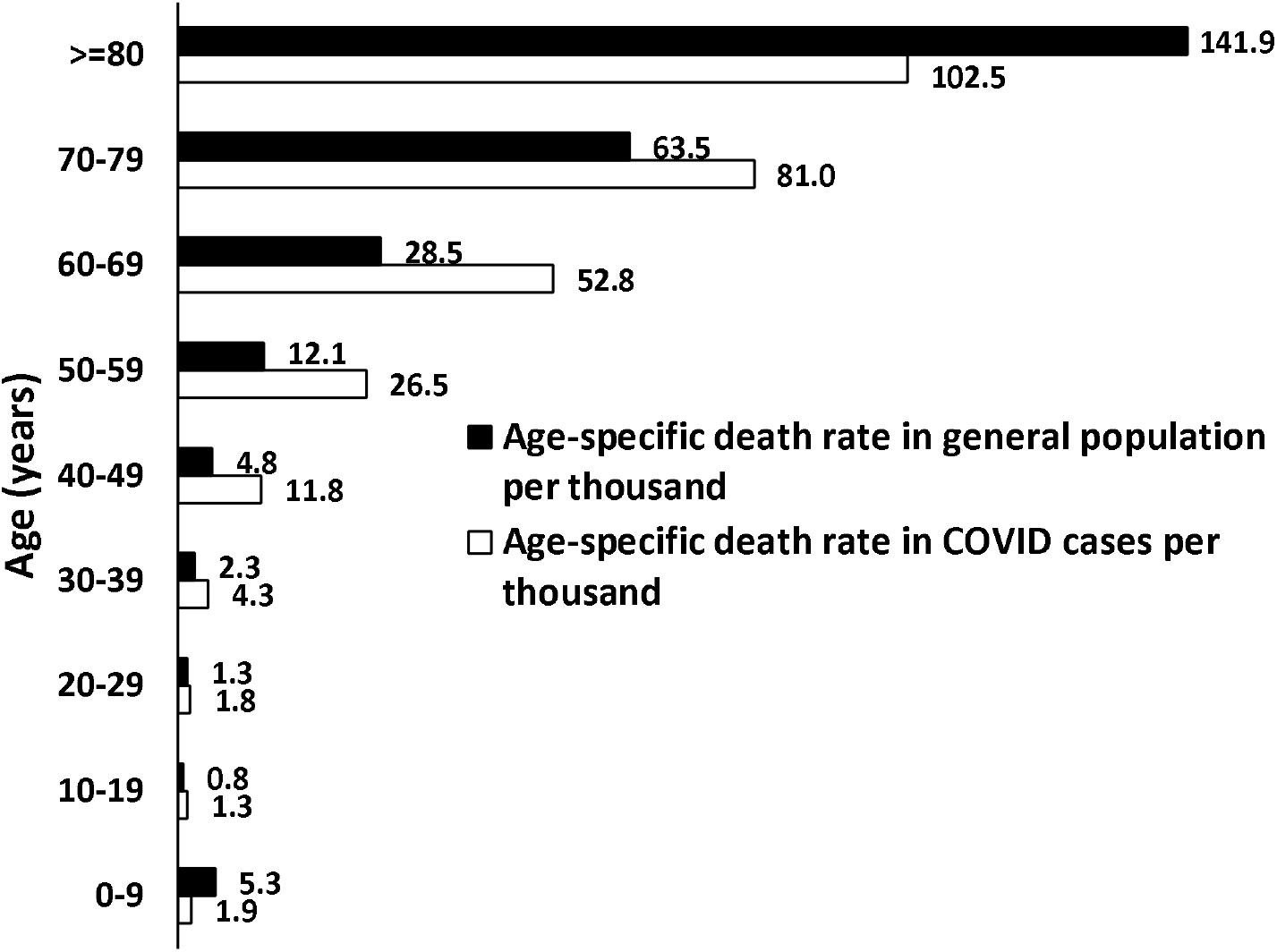
Age-specific death rate per 1000 cases of COVID cases and per 1000 population

When the expectation of life for deaths at different ages is used for calculating the life lost, the average YLL per COVID death in India is 19.90 years. Since mortality is 1.52%, the YLL=303 years per 1000 cases. In 7.2 million cases by October 17, 2020, this gives a total YLL 2.18 million. This is the years of life lost already due to this disease.

Considering the slow decline in the occurrence of new cases after reaching peak of 97,894 cases on September 17, 2020, and the likelihood of the second wave, the total cumulative cases in one-year may reach 13 million. This gives an incidence of nearly 1%. With this, total YLL in full year is expected to be 4.04 million. This comes to 3.026 YLL per 1000 population.

The total population of India as of October 2020 is estimated 1336 million and 9.6 million deaths expected during 2020 [7]. The age-distribution of deaths [6] gives an estimate of a total of nearly 231 million or 172 YLL per 1000 population during 2020 in India for all causes. Thus, YLL by COVID would be nearly 1.75% of the total YLL in India for all causes. The WHO estimate for total YLL for India in year 2016 [8] was nearly 359 million for all causes but this is based on the world’s highest projected expectation of life of 91.93 years. Our estimate of YLL based on India life table is nearly one-half of this.

The real brunt of COVID starts from the age of 20 years. When restricted to this age, the YLL by COVID is 331 years per 1000 cases and 3.306 per 1000 population of this age in full year. In the general population, this is 163 years per 1000 population for all causes. COVID YLL are nearly 2.0% of the total YLL from all causes for age 20 years and above.

A pointwise summary of these findings along with our comments is as follows:

1. YLL:
  a. Each COVID death in India on average is causing loss of nearly 20 years of life.
  b. Nearly 2.18 million years of life (YLL) have already lost in India by COVID deaths by the middle of October 2020. They are expected to exceed 4 million in a full year.
  c. YLL per 1000 cases of COVID could be nearly 303 years and nearly 3 per 1000 population in full year. This is just about 1.73% of the total YLL from all causes.
2. Cases:
  a. The share (14.5%) of age 50–59 years in COVID cases was nearly double of the share (7.7%) of this age-group in the general population. Thus, this is the age-group with highest relative burden of COVID disease in India. Contrary to belief, it is not the old age-group that has disproportionate vulnerability to this disease.
  b. The share (0.9%) of age-group 80+ years in COVID cases was nearly the same (0.8%) as in the general population. This may be because old age-group people exercise caution and do not go out as much. The age-group 50–59 years is the most affected possibly because this is the most active segment going outdoors.
3. Deaths:
  a. Share (5.8%) of the age-group 80+ in COVID deaths is nearly one-third of (15.7%) share of this age-group in deaths in general. The perception that this age-group carries more risk of death is not based on evidence. Perhaps our medical services are doing an excellent job of saving lives of old age people.
  b. The onslaught of mortality by COVID was particularly high in the 50-59 years that accounted for 25.3% share of COVID deaths against only 12.9% share in deaths in the general population. Is this because the age-group 50–59 years is affected much more than other age-groups (see point 2(a))? Not really, because the rate per 1000 cases also gives the same picture. The age-specific mortality in the age-group 50-59 years is much higher at 26.5 per 1000 in COVID cases against 12.1 per 1000 in the general population. Thus, the age group 50–59 years was not just more vulnerable to COVID in India but also had higher risk of death.
  c. Perhaps the age-specific death rate is the best measure of the burden of mortality in different age-groups. This was particularly higher in COVID cases of age 40-69 years relative to the rate in the general population.

## Discussion

To provide a perspective of the impact of COVID deaths, we compare this with road injuries. The WHO estimate based on the highest life expectancy for YLL by road injuries in the year 2016 in India was 15.39 million [8]. With the Indian life expectancy, these may be around one-half of this on the pattern of total YLL mentioned earlier. Thus, this was nearly 7.7 million. Compared to this, the 4.04 million YLL by COVID in the whole year is just about 57.71% of the YLL due to road injuries. Thus, the loss of years of life by COVID in full year is expected to be less than 60% of the loss by road injuries. Road injury deaths are more common in the younger age groups that makes the YLL high. This comparison with road injury deaths shows that the loss of life years by COVID may not be as high as is generally believed.

Korean study [9] carried out for cases and deaths occurring between January to April 2020 reported 4423 YLLs due to COVID per 100,000 persons or nearly 44 per 1000 population. This is exceedingly high relative to only nearly 3 per 1000 population for India. Nurchis et al. [10] reported disability-adjusted life years (DALYs) loss by COVID in Italy based on the data from February to May 2020. Their calculation of YLL was based on a standard life table. They reported total YLL but not per 1000 cases or per 1000 population. They reported that more than 99% of DALYs lost were due to YLL.

Arolas et al. [11] estimated 4.364 million YLL by June 13, 2020, for 42 countries, and the average loss of 14.5 years per death. They found YLL by COVID 2 to 6 times the median YLL due to seasonal influenza. Quast et al. [12] studied COVID cases from February 1 to July 11, 2020 and estimated “roughly” 1.2 million YLL due to COVID-19 deaths in the US. They used actuarial life expectancy table for 2017. Hanlon et al. [13] used standard WHO life tables and observed in April 2020 for Italy that YLL was 14 in men and 12 in women per COVID death. Our estimate for India is that on average COVID cases died nearly 20 years earlier than they would have naturally died. This disproves the belief that those dying were already close to the end of their life. Our loss in terms of years of life may be higher because the affliction was more in the age-group 50– 59 years where the remaining expectation of life is 21.8 years. Our study highlights the focus on relatively high burden of COVID, both incidence and deaths, in the age group age 50–59 years that seems to have not caught up any attention yet.

### Limitations

Our estimates are based on reported cases and deaths. Questions are sometimes raised about their undercount. Even if they are undercount, the YLL per death and YLL per 1000 cases may still be nearly correct unless the unreported cases and deaths belong to a specific (say younger) age. This is not likely to be so because any undercount will be for all age-groups. Nonetheless, more comprehensive, and standardized updated data are needed for more accurate calculation. Second, our estimates are based on national life expectancy. They lack international comparability but are more realistic in Indian context. Third, it can be argued that factors such as comorbidities should be considered, particularly because COVID is believed to be more fatal for people with comorbidities. Such ‘displaced mortality’ could result in lower YLL than our estimate. Fourth, in the absence of the data on the expectation of life in the year 2020, we have use this and other information for the latest year with available data, which is about two years old. These values may not have changed during this short period of two years.

## Supporting information

Figures 1S and 2S in the Supplement

## Data Availability

All data are taken from the public domain

## Key Messages

1. The age-group disproportionally affected by COVID in India is 50–59 years, both for getting the disease and for death. It is not old age (80+) as is generally believed. Our medical services seem to be doing an excellent job of saving life of old age people from this disease as the death rate by COVID in them has been lower than in the general population.
2. On average, nearly 20 years of life is lost by COVID deaths in India. This is consistent with point 1 since the expectation of life at age 55 years in India is nearly 21 years.
3. COVID deaths in India may cause a loss of a total of more than 4 million life years in full year but this impact is expected to be much less than by road injuries. The disease is not likely to cause as much loss of life as is generally feared.

## Acknowledgement

None

## References

1. National Centre for Disease Control. COVID19 Dashboard: Graphical illustration of data from COVID-19 cases in India. https://ncdc.gov.in/ – Accessed October 18, 2020.

2. Hindustan Times. 90% of those killed by Covid in India are older than 40, 69% are men. September 2, 2020. https://www.hindustantimes.com/india-news/90-of-those-killed-by-covid-in-india-are-older-than-40-69-are-men/story-glg0Ct4rHQ1YVvZgnckUcM.html – Accessed October 18, 2020.

3. The Wire. Nearly Half the People Who Have Died of COVID-19 in India Are Younger Than 60: October 14, 2020. https://science.thewire.in/health/india-covid-19-mortality-comorbidities-age-health-ministry/ – Accessed October 18, 2020.

4. SRS Based Abridged Life Tables, 2014-18. Office of the Registrar General, India: 2020. https://censusindia.gov.in/Vital_Statistics/SRS_Life_Table/SRS%20based%20Abridged%20Life%20Tables%202014-18.pdf

5. Indian Council of Medical Research, Public Health Foundation of India, and Institute for Health Metrics and Evaluation. India. Health of the Nation’s States - The India State-Level Disease Burden Initiative. New Delhi, India: ICMR, PHFI, and IHME 2017: p39. https://www.healthdata.org/sites/default/files/files/policy_report/2017/India_Health_of_the_Nation%27s_States_Report_2017.pdf

6. Sample Registration System Statistical Report 2018. Office of the Registrar General, India, 2020. https://censusindia.gov.in/Vital_Statistics/SRS_Report_2018/SRS_Statistical_Report_2018.pdf

7. Statista: India. Estimated total population from 2014 to 2024. https://www.statista.com/statistics/263766/total-population-of-india/ – Accessed October 21, 2020.

8. WHO. Health Statistics and Information Systems, Disease Burden and Mortality Estimates, Disease Burden 2000–2016, YLL estimates 2000–2016. https://www.who.int/healthinfo/global_burden_disease/estimates/en/index1.html – Accessed October 18, 2020.

9. Jo MW, Go DS, Kim R, et al. The burden of disease due to COVID-19 in Korea using disability-adjusted life years. J Korean Med Sci. 2020 Jun;35(21):e199. https://doi.org/10.3346/jkms.2020.35.e199

10. Nurchis MC, Pascucci D, Sapienza, M, et al. Impact of the burden of COVID-19 in Italy: Results of disability-adjusted life years (DALYs) and productivity loss. Int J Environ Res. Public Health 2020;17:4233.

11. Arolas HP i, Acosta E, Casasnovas GL, Lo A, Nicodemo C, Riffe T, et al. Global years of life lost to COVID-19. 2020 Jun 20: medRxiv 2020.06.19.20136069; doi: https://doi.org/10.1101/2020.06.19.20136069

12. Quast T, Andel R, Gregory S, Storch EA. Years of life lost associated with COVID-19 deaths in the United States. J Public Health (Oxf). 2020 Sep 7;fdaa159. doi: 10.1093/pubmed/fdaa159. Epub ahead of print.

13. Hanlon P, Chadwick F, Shah A, et al. COVID-19 – exploring the implications of long-term condition type and extent of multimorbidity on years of life lost: a modelling study [version 1; peer review: 1 approved]. Wellcome Open Res 2020;5:75. https://doi.org/10.12688/wellcomeopenres.15849.1

