## Supplementary material for "Preliminary Estimates of Years of Life Lost (YLL) Due to COVID-19 in India": Figures 1S and 2S in the Supplement

The following two figures provide visual comparison of the age distribution of COVID cases, COVID deaths, general population and deaths in the general population.

| Age (years) | Percent distribution of COVID cases | Percent distribution of general population |
| --- | --- | --- |
| 0-9 | 3.9% | 16.6% |
| 10-19 | 7.99% | 19.5% |
| 20-29 | 22.33% | 20.9% |
| 30-39 | 21.63% | 15.7% |
| 40-49 | 17.33% | 11.5% |
| 50-59 | 14.52% | 7.7% |
| 60-69 | 8.24% | 5.1% |
| 70-79 | 3.19% | 2.3% |
| >=80 | 0.86% | 0.8% |

Figure 1S. Age distribution of COVID cases in India in comparison to the age distribution in the general population

| Age (years) | Percent distribution of deaths to total deaths in COVID cases | Percent distribution of deaths to total deaths in general population |
| --- | --- | --- |
| 0-9 | 0.5% | 12.2% |
| 10-19 | 0.7% | 2.1% |
| 20-29 | 2.6% | 3.9% |
| 30-39 | 6.1% | 5.1% |
| 40-49 | 13.4% | 7.7% |
| 50-59 | 25.3% | 12.9% |
| 60-69 | 28.6% | 20.1% |
| 70-79 | 17% | 20.2% |
| >=80 | 5.8% | 15.7% |

Figure 2S. Age distribution of COVID deaths in India in comparison to the age distribution of all deaths in the general population
